# Validation of the youth version of the Alimetry® Gut-Brain Wellbeing Survey: A mental health scale for young people with chronic gastroduodenal symptoms

**DOI:** 10.1101/2025.06.05.25329082

**Authors:** Mikaela Law, Gayl Humphrey, Isabella Pickering, Gabriel Schamberg, Chris Varghese, Peng Du, Charlotte Daker, Hayat Mousa, Armen Gharibans, Greg O’Grady, Christopher N. Andrews, Stefan Calder

**Affiliations:** The Department of Surgery, The University of Auckland, New Zealand; Alimetry Ltd., Auckland, New Zealand; The Department of Psychological Medicine, The University of Auckland, New Zealand; Auckland Bioengineering Institute, The University of Auckland, New Zealand; The Department of Gastroenterology, Te Whatu Ora - Waitematā, New Zealand; Children’s Hospital of Philadelphia, Division of Gastroenterology, Philadelphia, Pennsylvania, United States of America; Perelman School of Medicine, University of Pennsylvania, Pennsylvania, United States of America; The Division of Gastroenterology, Cumming School of Medicine, University of Calgary, Canada

**Keywords:** youth, adolescents, anxiety, depression, stress, disorders of gut-brain interaction, psychometrics, mental health

## Abstract

**Background:** Chronic gastroduodenal symptoms present significant challenges for young people, often negatively impacting their quality of life and mental health. However, there is currently a lack of validated tools to assess mental health in young people with chronic gastroduodenal symptoms. This research outlines the development and validation of the Alimetry® Gut-Brain Wellbeing Survey-Youth Version (AGBW-Y), a novel tool for assessing mental health in young people with chronic gastroduodenal symptoms aged 12-17 years.

**Methods:** An iterative multi-phase approach was employed. In Phase 1, feedback was gathered from 19 paediatric clinicians in the gastroenterology field and 33 young people over multiple rounds to adapt the adult version of the AGBW Survey to be age-appropriate for young people aged 12-17 years. In Phase 2, rigorous psychometric testing of the adapted scale was conducted in a sample of 128 patients aged 12-17 years with chronic gastroduodenal symptoms, using an anonymous online survey.

**Results:** Based on the feedback from Phase 1, an interdisciplinary team of experts improved the survey’s language and usability for young people, including the removal of reverse-coded items. These adjustments enhanced the scale’s clarity, acceptability, and face and content validity. The final AGBW-Y comprises a patient preface, an opt-out option, 10 closed-ended questions, and an optional open-ended question. It assesses general mental health, alongside subscales of depression, stress, and anxiety. Phase 2 demonstrated excellent psychometric properties of the scale, including high internal consistency (α= .91 for the total scale; α= .75-.85 for subscales) and strong convergent, divergent, and concurrent validity with large effect sizes.

**Conclusions:** The AGBW-Y is a brief, reliable, and valid tool to assess mental health in young people with chronic gastroduodenal symptoms. This novel scale was developed through a rigorous co-design process with clinicians and young people, ensuring it is contextually relevant and clinically impactful. The AGBW-Y complements existing physiological assessments, enabling more accurate mental health evaluations which can be used to guide psychological referrals, support multidisciplinary care, and evaluate treatment outcomes.

## Introduction

Chronic gastroduodenal symptoms, particularly those associated with disorders of gut-brain interaction (DGBIs), are common and impactful in young people [1]. Within this population, the Rome IV criteria define several prevalent childhood gastroduodenal DGBIs, including cyclic vomiting syndrome, functional nausea, functional vomiting, rumination syndrome, aerophagia, and functional dyspepsia [2]. These conditions are characterized by a constellation of chronic and often bothersome symptoms such as nausea, vomiting, stomach pain, epigastric pain/burning, excessive fullness, early satiety, regurgitation, belching, and flatulence. These enduring gastroduodenal symptoms present significant challenges for young people and are associated with lower quality of life [3,4], poorer mental health outcomes [5,6], increased healthcare costs [7], and a greater frequency of medical visits [8,9]. Compounding these issues, the underlying pathophysiology of these disorders often remains elusive. This has consequently hindered the development of targeted therapy and has resulted in treatment strategies focused primarily on symptom management, without addressing underlying disease mechanisms.

An expanding body of research, guided by the biopsychosocial model, increasingly highlights the strong connection between gut health and mental wellbeing, particularly during the critical developmental stage of adolescence [10,11]. This relationship is bidirectional, whereby psychological comorbidities are frequently observed in young people with DGBIs [5,12], and conversely, feelings of stress, anxiety, and depression can trigger and exacerbate gastrointestinal symptoms [13–15]. This association is largely due to the gut-brain axis, a complex neurohormonal communication system between the central nervous system and the gastrointestinal tract [16–18]. Encouragingly, psychological interventions have demonstrated the potential to yield improvements in both mental health and gastrointestinal symptoms experienced by this patient population [11,19,20]. Recognizing this, psychological assessments have become an increasingly vital component of comprehensive clinical care for these patients [18,21–23]. Nevertheless, because the contributions of psychological factors and gastroduodenal symptoms are complexly intertwined, mental health issues in young people with childhood DBGIs often remain underdiagnosed and, as a result, undertreated. Therefore, the early identification and proactive addressing of mental health concerns emerge as crucial steps toward achieving improvements in the symptoms and quality of life for young individuals living with chronic gastroduodenal symptoms.

While the importance of the gut-brain connection and routine mental health assessment in patients with these symptoms is well-established [24], there is a notable lack of suitable clinical tools tailored specifically to this group [22]. Existing mental health scales are rarely designed for clinical application in patients with gastroduodenal symptoms, and are often inaccessible or developmentally inappropriate for young people. As a result, clinicians frequently rely on generalized mental health tools developed for adult populations, which inherently lack the necessary relevance and specificity for these younger patients. Adult screening tools often employ language and phrasing that are not developmentally aligned with the cognitive and emotional understanding of young people, potentially leading to misunderstandings and compromising the validity of the assessment when used with this age group [25]. Compounding the issue, many standard screening instruments for anxiety and depression include items that inquire about bodily symptoms. For young people with chronic gastroduodenal issues, these items may inadvertently overlap with their existing condition, making it difficult to distinguish between physical and psychological distress [22]. This overlap introduces the risk of inaccurate estimations of their true mental health status, raising fundamental questions around whether elevated scores are genuinely indicative of a mental health condition or merely a reflection of their underlying gastrointestinal symptoms.

Our team has previously developed and validated a novel mental health scale for patients with gastroduodenal symptoms, the Alimetry® Gut-Brain Wellbeing (AGBW) Survey [26]. The AGBW Survey was developed and validated using a patient-centred multi-phase process, described elsewhere [26]. The AGBW Survey comprises a patient preface, 10 closed-ended questions, and an optional open-ended question. This multidimensional scale assesses general mental health, alongside specific subscales relating to depression, stress, and anxiety, with increased specificity and contextualization tailored for use in patients with chronic gastroduodenal symptoms. However, the AGBW Survey was initially developed and validated for patients aged 18 years and over, raising the possibility that its language and content may not be optimally age-appropriate for young people.

Therefore, this research details the comprehensive development and validation of the Alimetry® Gut-Brain Wellbeing Survey-Youth Version (AGBW-Y), a novel tool designed to assess mental health in young people aged 12-17 years who experience chronic gastroduodenal symptoms.

## Methods

An iterative multi-phase approach was employed to develop and validate the AGBW-Y. Each phase involved co-design with paediatric gastroenterologists, GI psychologists, and young people aged 12-17 years to ensure face and content validity, comprehensibility, and acceptability. Ethical approval was received from the New Zealand Health and Disability Ethics Committee (2024 FULL 19553), and the trial was pre-registered at clinicaltrials.gov (NCT06394154).

### Phase 1. Adaptation and Feedback

The initial phase focused on evaluating the suitability of the existing adult version of the AGBW Survey for use in young people aged 12-17 years, specifically concerning its acceptability and validity within this younger demographic. To achieve this, a brief online scoping survey was sent to young people aged 12-17 years (with and without gastrodudoenal symptoms) and clinicians in the paediatric gastroenterology field. Both groups were presented with the adult version of the AGBW Survey. They were then asked a series of open-ended questions designed to explore the scale’s acceptability, simplicity, comprehensibility, and overall utility for patients in the 12-17 year age range. Furthermore, open-ended feedback was gathered on potential areas for improvement, seeking insights into how the scale could be modified to enhance its acceptability and ease of understanding for the intended youth audience.

The feedback gathered from this initial scoping survey then served as the foundation for adapting the language and structure of the original scale into a youth-specific version: the AGBW-Y. The adaptation process was undertaken by an interdisciplinary team of experts comprising two health psychology researchers specializing in psychogastroenterology, a gastroenterologist, a gastrointestinal surgeon, a paediatric digital health researcher, and two bioengineers specializing in gastric electrophysiology. Following the initial adaptation, the revised version was distributed to both the original cohort of clinicians and young people, as well as a new group of participants, to elicit further feedback to iteratively refine the scale. Multiple rounds of feedback were conducted until a consensus of satisfaction with the final version of the youth scale was reached among both clinicians and youth. This refined the acceptability, clarity, comprehensibility, and face and content validity of the scale for patients aged 12-17 years with chronic gastroduodenal symptoms.

### Phase 2. Psychometric Validation

The psychometric properties of the scale were assessed using an anonymous, cross-sectional survey involving patients aged 12-17 years with chronic gastroduodenal symptoms. Informed consent was obtained from all participants.

#### Sample

Patients were recruited through convenience sampling using social media, flyers, and clinic email lists. Patients were included if they were between 12 and 17 years old and fluent in English. They also needed to meet the Rome IV symptom criteria [2] and/or have a self-reported previous clinical diagnosis for at least one of the following childhood gastroduodenal DGBIs: gastroparesis, cyclic vomiting syndrome, functional nausea, functional vomiting, rumination syndrome, aerophagia, or functional dyspepsia. Individuals with self-induced vomiting or self-reported eating disorders were excluded. Recruitment efforts were conducted globally, focusing on ensuring a diverse representation across geographic regions, ages, and genders. A sample of at least 100 young people was recruited, due to recommendations that a validation sample should consist of at least 10 respondents per scale item [27].

#### Procedure

The anonymous survey was conducted online using Qualtrics (Qualtrics, Provo, UT) and took approximately 10-15 minutes to complete. All participants provided informed consent based on previously validated consent processes for internet-based research with young people [28,29]. Participants were asked to read a set of consent clauses and check a box to indicate their consent. Respondent’s capacity and maturity to provide informed consent were then assessed using two multiple-choice questions about the consent process. If a participant incorrectly answered both of these questions after two attempts, they could not continue with the study.

After demonstrating informed consent, participants were presented with a demographics questionnaire, followed by a battery of youth psychological questionnaires, including the AGBW-Y, presented in a randomized order. The survey concluded with an optional feedback form regarding the scale, and participants had the opportunity to enter a prize draw to thank them for their responses. Responses were collected between July 2024 and April 2025.

#### Measures

##### Psychometrics

The following psychometrics were measured to assess convergent validity: the Patient Health Questionnaire for Adolescents (PHQ-A) [30] to measure depression, the Generalized Anxiety Disorder 7 (GAD-7) [31,32] to measure anxiety, the PROMIS Pediatric Psychological Stress Experiences Short Form 8a (PPSE) to measure psychological stress [33], the Depression Anxiety and Stress Scale Youth Version (DASS-Y) [34] to measure anxiety, depression, and stress in an integrated scale, and the Kessler Psychological Distress Scale (K10) [35] to measure total levels of distress and mental health.

The Big Five Questionnaire for Children (BFQ-C) energy/extraversion subscale [36], which measures levels of extraversion in childhood and adolescence, and the Emotion Regulation Questionnaire for Children and Adolescents (ERQ-CA) [37], which measures emotion regulation in children and adolescents, were used to assess divergent validity. Lastly, concurrent validity was assessed using the Pediatric Quality of Life Inventory 4.0 (PedsQL) [38], which measures health-related quality of life in children and young people.

All psychometrics used for validity testing have been validated for use in young people aged 12-17 years.

##### AGBW-Y feedback form

To gather insights into the user experience, an optional feedback form was presented at the end of the survey. This form asked respondents to evaluate the AGBW-Y using a visual analogue scale for two attributes; 1) how easy the questionnaire is to complete on a scale of 0 (very hard) to 100 (very easy) and 2) how easy the questions are to understand on a scale of 0 (very hard) to 100 (very easy). Additionally, they were invited to share any additional comments or suggestions regarding the AGBW-Y.

#### Statistical Analysis

Data were analyzed using IBM SPSS Statistics v29. A *p*-value of .05 was considered statistically significant. To maximize data inclusion, partial responses were included, provided patients completed the demographic information and the first four questions of the AGBW-Y (depression subscale). This resulted in a variable number of respondents for validity and reliability analyses, but each calculation included at least 119 participants.

##### Reliability

Internal consistency reliability of the subscale and total scores was examined using Cronbach’s alpha coefficients (α), with a value of α> .70 indicating acceptable reliability, α> .80 ideal reliability, and α> .90 excellent reliability [27,39–41]. To assess the consistency between scale items and their relationship with the subscale/total scores, inter-item correlations and corrected item-total correlations were calculated, with an *r*>.30 indicating good consistency [27,40].

##### Validity

To establish sufficient construct and criterion validity for the subscale and total scores, at least 75% of the predefined hypotheses for convergent, divergent, concurrent, and known-groups validity were required to be supported for each subscale/total score [41,42]. Pearson’s correlation coefficients assessed convergent, divergent, and concurrent validity. A correlation coefficient *r*> .50 indicated good convergent validity [40,42,43]. Divergent validity was considered successful if the correlations were weaker than those observed for convergent validity [40,44]. Positive and statistically significant correlations were expected for concurrent validity, with *r*> .50 indicating strong evidence of concurrent validity. To assess known groups validity, one-tailed independent samples t-tests were performed, testing the hypothesis that females were expected to score significantly higher than males on the subscale and total scores [45–48].

## Results

### Phase 1. Adaptation and Feedback

#### Sample

A total of 33 young people aged 12-17 years and 19 paediatric clinicians provided feedback on the scale. The clinician sample consisted of 11 gastroenterologists, four dieticians, one surgeon, one psychologist, one nurse, and one hospital medicine specialist, from the USA, New Zealand, Australia, and Canada, all specialising in paediatrics. The youth sample was recruited globally and included 17 females, 14 males, and two gender diverse individuals, with an average age of 14.42 years (*SD=* 1.60).

#### Feedback rounds

Three rounds of feedback were gathered and used to iteratively adapt the scale. The initial round confirmed that the current adult version of the AGBW Survey was inappropriate for use in patients aged 12-17 years. In response, the survey’s language and usability were modified for young people, including removing the two reverse-coded items from the stress subscale. These adjustments successfully enhanced the scale’s clarity, acceptability, comprehensibility, and face and validity, whilst preserving the original constructs within each question. The final version of the scale was perceived as user-friendly and easy to understand.

### The Final Scale: The Alimetry® Gut-Brain Wellbeing Survey- Youth Version (AGBW-Y)

The final AGBW-Y (see Supplementary File) consists of a patient preface, 10 closed-ended questions, and a final optional open-ended question. The patient preface explains the purpose of these questions and how the information will be used. The 10 closed-ended questions ask patients to rate how often they have felt or behaved in a certain way over the last two weeks on a scale of 0 (none of the time) to 4 (all of the time). The 10 questions can be summed to create a total score (maximum 40). Additionally, three subscale scores can also be calculated for: depression (sum of the first four questions, maximum 16), stress (sum of the middle three questions, maximum 12), and anxiety (sum of the last three questions, maximum 12). Higher scores indicate worse mental health. The scale ends with an optional free-text question for patients to provide additional comments about their wellbeing.

### Phase 2. Psychometric Validation

#### Sample Characteristics

A sample of 128 young people completed the validation survey (mean age= 15.48 years, *SD*= 1.60). The majority of respondents were female, white, and from the USA or British Commonwealth (Table 1). A high degree of comorbidity was observed among Rome IV diagnoses, with many patients meeting criteria for multiple conditions, particularly functional dyspepsia, functional nausea, and aerophagia. Only 15% met criteria for a single gastroduodenal DGBI, and over half (n=65, 51%) met criteria for three or more.

**Table 1.**
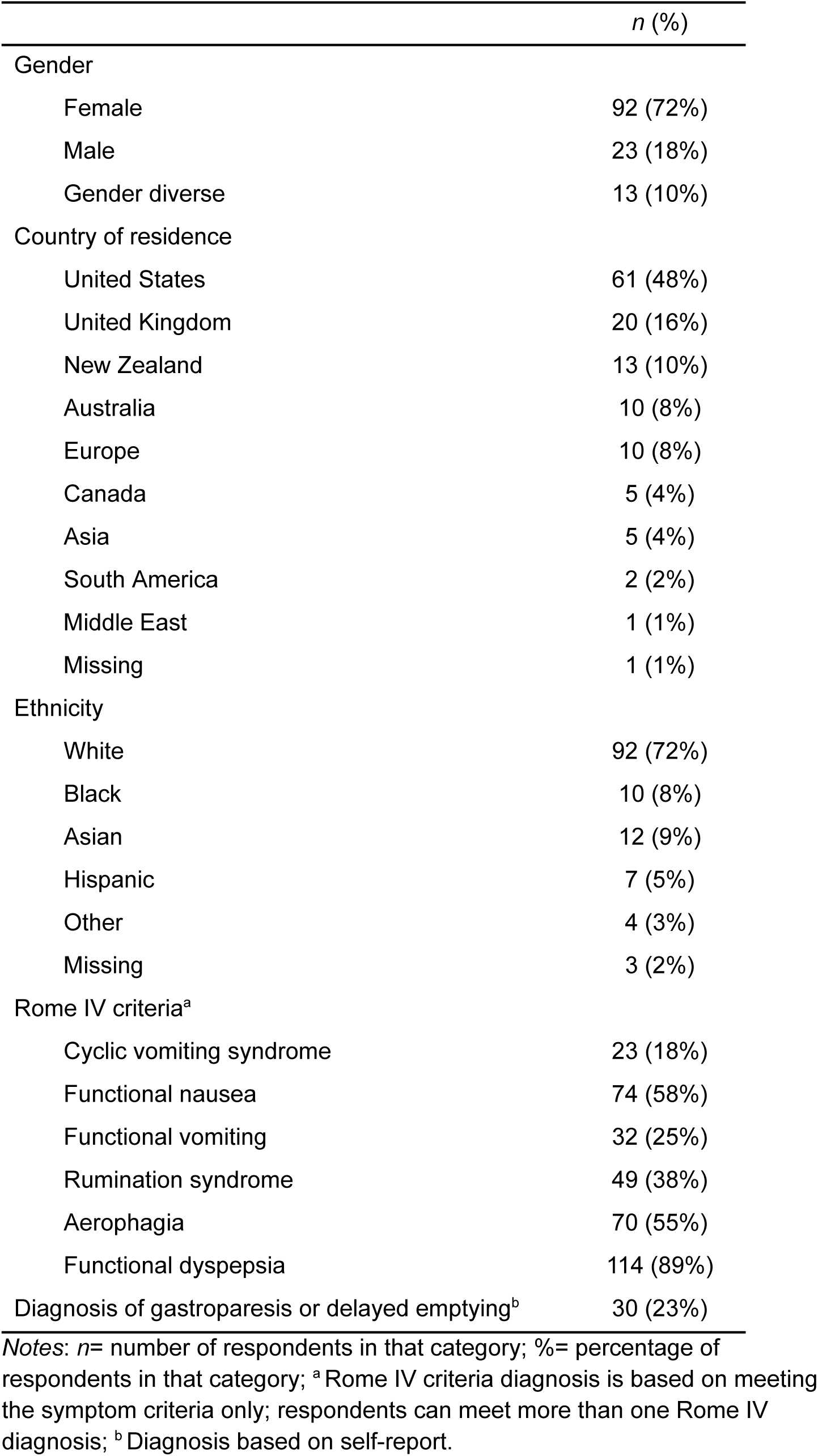
Demographic characteristics of the survey respondents (*N*= 128)

#### Descriptive Statistics

The 10 questions took participants a median of 56 seconds to complete (*IQR*= 45-76 seconds). Table 2 indicates that the full range of subscale scores was used, with all individual questions receiving responses across the entire 0-4 range. As evidenced by the skewness and kurtosis values in Table 2 and the distribution graphs in Figure 1, all subscale and total scores were normally distributed. The mean scores, which fell within the middle of the scale range, further support this normality.

**Figure 1.**
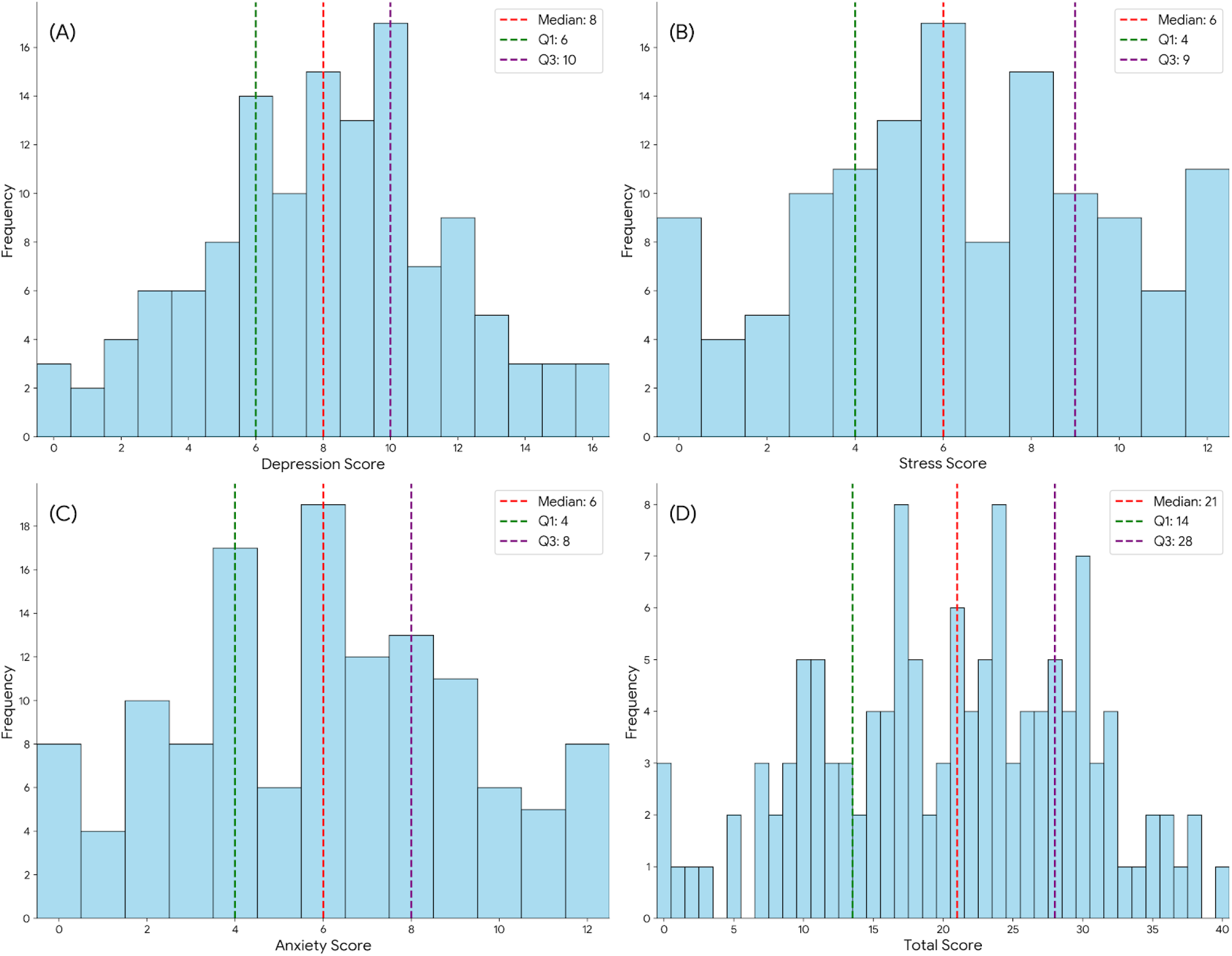
Distribution of the AGBW-Y subscale and total scores. Histograms depict (A) depression, (B) stress, and (C) anxiety subscale scores, and (D) the total score.

**Table 2.** Descriptive statistics of the AGBW-Y Survey’s subscales and total scores.

| Scale | *N* | *M* | *SD* | Range | Skewness | Kurtosis | $\alpha$ |
| --- | --- | --- | --- | --- | --- | --- | --- |
| Depression subscale | 128 | 8.11 | 3.63 | 0-16 | -0.06 | -0.32 | .75 |
| Stress subscale | 128 | 6.32 | 3.42 | 0-12 | -0.09 | -0.81 | .85 |
| Anxiety subscale | 127 | 5.97 | 3.29 | 0-12 | 0.12 | -0.77 | .84 |
| Total score | 127 | 20.41 | 9.30 | 0-40 | -0.18 | -0.61 | .91 |

Notes: *N*= number of respondents; *M*= mean; *SD*= standard deviation; $\alpha$ = Cronbach's alpha coefficient.

#### Reliability

Table 2 demonstrates strong reliability for the AGBW-Y. The total score showed excellent internal consistency reliability, while the anxiety and stress subscales demonstrated good reliability, and the depression subscale, acceptable reliability. Removing items did not result in any increases in reliability. Inter-item correlations (r = .37-.70) and corrected item-total correlations (r = .48-.80) indicated good consistency between the individual items and the items and subscale and total scores, respectively.

#### Validity

##### Convergent validity

Good convergent validity was demonstrated by significant correlations (all *p*s<.001) with large effect sizes observed between the AGBW-Y’s subscale and total scores and the other mental health scales measured (Table 3).

**Table 3.** Pearson correlation coefficients between the AGBW-Y Survey scores and comparative questionnaires used for validity testing.

|  |  | Depression subscale | Stress subscale | Anxiety subscale | Total score |
| --- | --- | --- | --- | --- | --- |
| Convergent validity | PHQ-A total | .84* |  |  |  |
|  | DASS-Y depression subscale | .77* |  |  |  |
|  | PPSE total |  | .75* |  |  |
|  | DASS-Y stress subscale |  | .53* |  |  |
|  | GAD-7 total |  |  | .87* |  |
|  | DASS-Y anxiety subscale |  |  | .72* |  |
|  | K-10 total |  |  |  | .85* |
|  | DASS-Y total |  |  |  | .84* |
| Divergent validity | BFQ-C energy/extraversion subscale | -.35* | -.21* | -.19* | -.28* |
|  | ERQ-CA cognitive reappraisal subscale | -.23* | -.19* | -.19* | -.23* |
|  | ERQ-CA expressive suppression subscale | .38* | .32* | .34* | .39* |
| Concurrent validity | PedsQL total | -.61* | -.62* | -.51* | -.65* |
|  | PedsQL emotional subscale | -.63* | -.70* | -.81* | -.79* |

*Notes:* AGBW-Y= Youth Version of the Alimetry® Gut-Brain Wellbeing Survey; PHQ-A= Patient Health Questionnaire for Adolescents; DASS-Y= Depression Anxiety and Stress Scale Youth Version; PPSE= PROMIS Pediatric Psychological Stress Experiences Short Form 8a; GAD-7= Generalized Anxiety Disorder 7; K-10= Kessler Psychological Distress Scale; BFQ-CA= Big Five Questionnaire for Children; ERQ-CA= Emotion Regulation Questionnaire for Children and Adolescents; PedsQL= Pediatric Quality of Life Inventory 4.0; \* denotes significance at $p < .05$ .

##### Divergent validity

The AGBW-Y subscale and total scores showed small to moderate correlations with the BFQ-C and ERQ scores (Table 3). As shown in Table 3, these correlations were substantially weaker than those observed for convergent validity correlations, indicating successful divergent validity.

##### Concurrent validity

Concurrent validity was demonstrated by significant correlations (all *p*s<.001) with large effect sizes observed between the AGBW-Y subscales and total score and the PedsQl total score and emotional subscale score (Table 3).

##### Known groups validity

As shown in Table 4, there were no significant differences between males and females on any of the subscale or total scores. However, females had higher average scores than males on all subscales.

**Table 4.** Results from the independent samples t-tests used for known groups validity of the AGBW Survey.

|  |  | N | Depression subscale | Stress subscale | Anxiety subscale | Total score |
| --- | --- | --- | --- | --- | --- | --- |
| Gender | Female, M (SD) | 92 | 8.41 (3.67) | 6.35 (3.44) | 5.88 (3.17) | 20.64 (9.20) |
|  | Male, M (SD) | 23 | 7.26 (3.72) | 5.30 (3.24) | 5.50 (3.61) | 18.09 (10.16) |
|  | *p*-value |  | .091 | .095 | .312 | .127 |
|  | Cohen's *d* |  | 0.31 | 0.31 | 0.12 | 0.27 |

Notes: M= mean; SD= standard deviation; *p*-values were calculated using independent samples t-tests; \* denotes significance at $p < .05$ .

#### Feedback Form

The optional feedback form was completed by 107 patients, who rated the ABGW-Y as easy to complete (*M*= 80.69, *SD*= 17.40) and easy to understand (*M*= 83.62, *SD*= 18.80) on the 100-point visual analogue scales. A smaller group of 23 patients also provided qualitative feedback, highlighting an appreciation for the simple wording and overall ease of understanding. The final open-ended question was well-received as patients valued the opportunity to elaborate on their responses, such as whether their experiences felt ‘normal’ and their perspectives on the cause-and-effect relationship between their mental and physical health. Importantly, patients also expressed their gratitude for this questionnaire, describing it as a valuable outlet to share their experiences, particularly during challenging times, such as following a recent diagnosis that had significantly impacted their mental wellbeing.

## Discussion

This research details the systematic development and validation of the AGBW-Y, a novel instrument specifically tailored for the assessment of mental health in young people aged 12-17 years who experience chronic gastroduodenal symptoms. The AGBW-Y offers a rapid and efficient means of assessing mental health by integrating evaluations of depression, stress, and anxiety that are specifically contextualized for the unique experiences of patients with chronic gastroduodenal symptoms.

Employing a comprehensive multi-phase, mixed-methods approach that incorporated co-design with young people and paediatric clinicians, we developed a tool that addresses a significant gap in clinical practice - the lack of age-appropriate mental health assessments for this vulnerable patient group. Many widely used mental health assessments include questions pertaining to physical symptoms, such as changes in appetite or sleep disturbances, which may be directly attributable to their underlying gastrointestinal condition, rather than solely reflecting psychological factors [22]. By contextualizing mental health assessment specifically for this population, the AGBW-Y offers a valid and accurate reflection of mental health, thereby potentially reducing the overestimation of psychological concerns that can occur when employing generic assessment tools.

The AGBW-Y provides a brief yet comprehensive multidimensional assessment of key psychological constructs, namely depression, stress, and anxiety. This single, efficient scale allows for a rapid understanding of how a young patient is feeling without imposing the burden of multiple extensive evaluations for both the young person and the clinician. The simplified language and more straightforward structure of the AGBW-Y contribute to its enhanced accessibility for young people, all while ensuring the preservation of the core psychological constructs included in the adult version. Importantly, the AGBW-Y was perceived by young people as easy to complete and understand, suggesting a high level of acceptability and feasibility for integration into clinical settings. The inclusion of the optional open-ended question was particularly appreciated by patients, as it provided a valuable opportunity to offer additional context to their quantitative responses and communicate any further concerns directly to their healthcare providers.

One key adaptation from the adult version was the deliberate removal of the reverse-coded items in the stress subscale. This modification was directly informed by the feedback process and enhanced the clarity and overall usability of the tool. This aligns with previous research indicating that reverse-coded items can inadvertently reduce the reliability of assessments by introducing systematic measurement error, particularly in younger populations where the nuanced understanding of negatively worded items is often less developed [49–51]. Indeed, the internal consistency reliability of the stress subscale in the AGBW-Y was demonstrably higher than that observed in the adult AGBW Survey [26], which does include reverse-coded items, providing further evidence of the increased ease of comprehension afforded by this change.

Furthermore, the AGBW-Y demonstrated robust psychometric properties, exhibiting high internal consistency reliability and strong evidence of convergent, divergent, and concurrent validity. These findings collectively indicate that the AGBW-Y effectively and accurately measures the intended psychological constructs within young people with chronic gastroduodenal symptoms. Notably, the average scores and validity results were comparable to those observed in the validation of the adult AGBW Survey [26], suggesting that the core constructs being measured have remained consistent and the scales provide comparable assessments across the two age groups. Similar to the adult AGBW Survey, the distribution of scores in the youth sample exhibited a normal pattern, characterized by minimal skew and kurtosis. This observation contrasts with the typically positively skewed distribution often seen in general population mental health measures [52,53] and likely reflects the higher prevalence of psychological comorbidities reported in young people with chronic gastroduodenal symptoms [5,6,12]. This further emphasizes the clinical significance of routine mental health assessment in this population.

Similar to the adult AGBW Survey, the AGBW-Y did not demonstrate a statistically significant ability to discriminate between male and female participants, contrary to our initial hypothesis, which is likely attributed to the low number of males recruited for this study. Despite targeted recruitment strategies, the recruitment of males proved to be a persistent challenge within this study, an issue commonly experienced in internet-based research studies [54,55]. However, when compared to the adult AGBW Survey validation [26], the AGBW-Y did show clearly divergent mean scores between males and female participants in the anticipated direction, along with trending *p*-values for the depression and stress subscales. This suggests that with a larger and more representative sample of males, this difference may indeed reach statistical significance. Future research should prioritize recruitment strategies specifically designed to engage young males to ensure their accurate representation within mental health research.

Research into the mental health of young people with chronic gastroduodenal symptoms is often overlooked. However, adolescence represents a unique developmental period in a patient’s life, offering an opportune time to begin instilling essential coping skills that can serve them as they mature. Therefore, the clinical implementation of the AGBW-Y holds several potential benefits, particularly when integrated alongside routine medical tests, such as body surface gastric mapping, to facilitate more holistic and comprehensive assessments. Firstly, the AGBW-Y can help identify young people who may benefit from targeted psychological support, thereby enabling early intervention strategies that have the potential to improve both their mental health and gastrointestinal symptoms [11,19,20]. Secondly, it can inform the development of more personalized and multidisciplinary treatment approaches, which have consistently demonstrated improved outcomes in this patient population [56,57]. Finally, the AGBW-Y can serve as a valuable outcome measure for evaluating treatment effectiveness both clinically and in research trials, particularly for interventions targeting the gut-brain connection. However, it is important to reiterate that while the AGBW-Y is a useful and reliable assessment tool, it is not intended to serve as a diagnostic instrument. Consequently, any suspected mental health issue necessitates a timely and appropriate referral to psychological services for a comprehensive evaluation and the development of an appropriate treatment plan.

The development and validation of the AGBW-Y within a diverse international sample significantly enhances its generalizability across different healthcare contexts and settings. However, it is important to acknowledge the predominance of female, white, and Western participants, which may limit applicability to all demographic groups. Future research should examine the scale’s performance across more diverse populations, particularly among different cultural contexts and ethnicities. Additionally, the AGBW-Y is currently available only in English, highlighting the need for rigorously validated translations to facilitate its use in other languages. Furthermore, given the cross-sectional design, we were unable to assess the scale’s predictive validity or responsiveness to change over time. Longitudinal research is warranted to determine how AGBW-Y scores relate to relevant clinical outcomes and treatment response. Lastly, while the scale was developed with input from a diverse range of clinicians, future research should investigate the practical effectiveness of the AGBW-Y in informing clinical decision-making within routine healthcare settings.

### Summary

The AGBW-Y is a brief, valid and reliable tool that represents a significant advancement in the assessment of mental health in young people with chronic gastroduodenal symptoms. By directly addressing a critical gap in both clinical practice and research, the AGBW-Y offers a much-needed age-appropriate measure for these patients. The co-design process, which actively involved both clinicians and young people, has ensured that the scale is contextually relevant, well-accepted by its intended users, and possesses significant clinical impact. The integration of the AGBW-Y into routine clinical assessments, alongside existing physiological evaluations, has the potential to support a more holistic and comprehensive approach to the care of these young patients, ultimately enabling more accurate evaluations of their mental health and potentially leading to improved overall outcomes for this vulnerable population.

## Data Availability

Data is available upon reasonable request to the corresponding author.

